# Systematic Review of risk score prediction models using maternal characteristics with and without biomarkers for the prediction of GDM

**DOI:** 10.1101/2023.10.23.23297401

**Authors:** Durga Parkhi, Swetha Sampathkumar, Yonas Weldeselassie, Nithya Sukumar, Ponnusamy Saravanan

## Abstract

**Background:** GDM is associated with adverse maternal and fetal complications. By the time GDM is diagnosed, continuous exposure to the hyperglycaemic intrauterine environment can adversely affect the fetus. Hence, early pregnancy prediction of GDM is important.

**Aim:** To systematically evaluate whether composite risk score prediction models can accurately predict GDM in early pregnancy.

**Method:** Systematic review of observational studies involving pregnant women of <20 weeks of gestation was carried out. The search involved various databases, grey literature, and reference lists till August 2022. The primary outcome was the predictive performance of the models in terms of the AUC, for <14 weeks and 14-20 weeks of gestation.

**Results:** Sixty-seven articles for <14 weeks and 22 for 14-20 weeks of gestation were included (initial search - 4542). The sample size ranged from 42 to 1,160,933. The studies were from Canada, USA, UK, Europe, Israel, Iran, China, Taiwan, South Korea, South Africa, Australia, Singapore, and Thailand. For <14 weeks, the AUC ranges were 0.59-0.88 and 0.53-0.95, respectively for models that used only maternal characteristics and for those that included biomarkers. For 14-20 weeks these AUCs were 0.68-0.71 and 0.65-0.92. Age, ethnicity, BMI, family history of diabetes, and prior GDM were the 5 most commonly used risk factors. The addition of systolic BP improved performance in some models. Triglycerides, PAPP-A, and lipocalin- 2, combined with maternal characteristics, have the highest predictive performance. AUC varied according to the population studied. Pooled analyses were not done due to high heterogeneity.

**Conclusion:** Accurate GDM risk prediction may be possible if common risk factors are combined with biomarkers. However, more research is needed in populations of high GDM risk. Artificial Intelligence-based risk prediction models that incorporate fetal biometry data may improve accuracy.

## Introduction

Gestational Diabetes Mellitus (GDM) is defined as any degree of glucose intolerance with onset or first recognition during pregnancy Rodriguez and Mahdy (2019). GDM is associated with many short-term and long-term complications for the mother and her offspring Plant et al. (2020). Women with GDM have a higher risk of pre-eclampsia and a higher risk of Type 2 diabetes or cardiovascular disorders (CVD) in the long term. Offspring born to women with GDM can have a higher risk of both short-term and long-term adverse outcomes including cardiometabolic disorders in the long term Saravanan et al. (2020). GDM is highly prevalent among South Asian women compared to women from other ethnic groups K. W. Lee et al. (2018). Some estimates suggest this may affect 1 in 5 pregnancies in India and the risk of type 2 diabetes in this population is higher than in other populations Shivashri et al. (2022).

There is increasing evidence that GDM is diagnosed too late in pregnancy (Savio et al, Diabetes care) and excess fetal adiposity may be present several weeks before the onset of GDM Sukumar et al. (2016). In addition, despite good control of GDM, adverse metabolic programming may exist with resultant accelerated adiposity accumulation in infants born to mothers with GDM Logan et al. (2016). Therefore, it is imperative not only to diagnose GDM early in pregnancy but also to prevent the onset of GDM in women who are at higher risk. Accurate prediction of incident GDM may enable prevention. FIGO Best Practice Advice recommends viewing pregnancy as an opportunity to prevent type 2 diabetes mellitus Adam et al. (2023).

Screening for GDM varies across countries. Universal screening is recommended in populations with a high prevalence of GDM and selective screening where the prevalence is low. However, by either strategy, the pickup rate is only between 5 and 15%. Hence many women undergo unnecessary testing. Currently, the common risk factors used for selective screening include age, BMI, family history (1st-degree relative) of diabetes, polycystic ovarian syndrome, previous history of GDM, previous history of macrosomia, and women from certain high-risk ethnic groups K. W. Lee et al. (2018), Pons, Rockett, de Almeida Rubin, Oppermann, and Bosa (2015). Different guidelines use different risk factors and none use composite risk scores. In addition to these ‘traditional’ risk factors, several other risk factors have been identified recently (such as triglycerides, adiponectin, the fetal abdominal wall thickness in early pregnancy, etc) but none have been included in any of the screening guidelines. Weighted incorporation of these risk factors in novel prediction methods using machine learning may also improve the predictive ability of GDM. Recently, several studies published the utility of such prediction models with mixed results (here cite the 2022 studies). We, therefore, conducted a systematic review of all the available studies that used composite risk prediction models before 20 weeks of gestation for the prediction of GDM. If proven, the accurate prediction may aid in implementing GDM prevention strategies early in pregnancy.

## Methods

### Research Design

This study was conducted according to PRISMA (Preferred Reporting Items for Systematic Reviews and Meta-Analyses) guidelines.

### Search Methods

We searched Medline, Embase, Cinahl, Web of Science, Scopus, and ProQuest databases, Cochrane trials, grey literature, and reference lists of relevant papers for articles published in English till August 2022. The main keywords for the search strategy included “gestational diabetes mellitus”, “pregnancy”, “glucose intolerance”, “hyperglyc*”, “prediction model”, “statistical model”, “machine learning”, “risk score”, and “risk assessment”. The detailed search criteria are listed in the Appendix. Additionally, the reference list of each identified study was manually searched to identify any additional studies. EndNote X7 was employed to manage the studies and remove duplicate items. The search was updated to include any additional studies on 28 Feb 2023.

### Inclusion and Exclusion Criteria

All included studies had to meet the following criteria: (1) published in English; (2) included pregnant women from the general population, with a clear definition for GDM diagnosis; (3) included composite risk score models for GDM prediction, with a clear description of the models and the time of collection of the predictors; and (4) showed the performance of the models in terms of auROC. Articles in other languages, other types of articles (eg, reports and reviews), or those that used other measures for GDM detection were excluded. Articles with prediction models using predictors collected at > 20 weeks of pregnancy or unknown time points were also excluded. Search and data extraction were performed independently by two investigators (DP and SS). Disagreements were resolved by discussion with a third reviewer (PS).

### Data Extraction

Data extraction was carried out by the Transparent Reporting of a Multivariable Prediction Model for Individual Prognosis or Diagnosis (TRIPOD) protocol Moons et al. (2015). The following data were extracted from each study: (1) demographic information (ie, the country in which the data were gathered, the setting, the study design, the prediction temporality, and the outcome definition); (2) the candidate and final features of the model training, the prediction model type (Regression/Bayesian/ML/Other), and the model validation and application (risk equation and cut-off, if given); (3) prediction outcomes, performance measures including auROC, sensitivity, specificity, ppv, and npv, and calibration, if available.

### Quality and Bias Assessments

The PROBAST questionnaire, which includes a total of 20 signaling questions in four domains (ie, participants, predictors, outcome, and analysis), was used as a tool for assessing the risk of bias and applicability of each included study Wolff et al. (2019).

### Statistical Analysis

The performance of each composite risk score (CRS) model was described using the primary outcome measures of discrimination and calibration. Model discrimination or concordance index (C-index) is similar to the AUROC Steyerberg et al. (2010) and indicates its diagnostic or prognostic discrimination ability as none (AUROC <=0.6), poor (AUROC >0.6 to 0.7), fair (AUROC >0.7 to 0.8), good (AUROC >0.8 to 0.9), or optimum (AUROC >0.9 to 1). Model calibration is a metric of the goodness of fit that assesses the agreement between observed and predicted outcomes and reflects the stability of the model via calibration plots. The *I*^2^ test was used to assess the statistical heterogeneity among the included studies Higgins et al. (2019). Where possible, the pooled estimates of AUROC, sensitivity, and specificity were calculated, but in presence of high heterogeneity between different CRS models, these were not carried out. The analysis of the included studies was divided into primary and subgroup analyses to judge the performances of the CRS methods in predicting GDM in different gestational period. Sensitivity analyses and subgroup analyses were also conducted to gain insight into potential sources of interstudy heterogeneity due to selector or inclusion criteria bias. The abilities of the different ML algorithms (eg, LR, Bayesian model, SVM, and XGBoost) for predicting GDM were also carried out as additional analysis.

## Results

### Study selection

A total of 123 studies were included out of the initial 4,542 studies. Out of these, 89 (72.36%) were in the gestational age range of less than 14 weeks or 14-20 weeks. Figure 1 shows the PRISMA flow diagram of the study selection process.

### Study characteristics

Among the 89 included studies, 67 used prediction models <14 weeks gestation (Table 2) and 22 between 14 and 20 weeks of gestation (Table 3). Majority of these studies were published within the last 10 year and 11 were published in 2022. Eleven used ML-based techniques and others mostly used regression or Bayesian methods for prediction. Most regression-based studies used either forward stepwise selection or backward stepwise selection for multivariable feature selection. The studies were from the USA, Canada, UK, Europe, Israel, Iran, China, Taiwan, South Korea, South Africa, Australia, Singapore, and Thailand. Despite >90% of GDM is estimated to occur in South and Southeast Asia very few studies were found. **Studies of <14 weeks gestation:** Sample size of these studies varied between xx and yy. Out of the 67 studies, eight used ML-based techniques (Figure 1). The predictive value of AUROC ranged from 0.59 to 0.88 for the models with only maternal characteristics and from 0.53 to 0.95 when the biomarkers were included. **Studies of 14-20 weeks of gestation:** Sample size of these studies varied between xx and yy. Out of the 22 studies, three used ML-based techniques (Figure 1). The predictive value of AUROC ranged from 0.68-0.71 for the models with only maternal characteristics and from 0.65-0.92 when the biomarkers were included.

**Table 1.**

| Sr. No. | Parameter | <14 weeks |  | 14-20 weeks |  |
| --- | --- | --- | --- | --- | --- |
|  |  | M | M+B | M | M+B |
| 1 | No. of studies | 21 | 46 | 3 | 22 |
| 2 | ML-based | 2 | 6 | 0 | 3 |
| 3 | Sample size range | 134-1,160,933 | 42-1,160,933 | 1303-105,379 | 103-417,601 |
| 4 | auROC range | 0.59-0.88 | 0.531-0.951 | 0.68-0.714 | 0.649-0.92 |
| 5 | Countries studied | USA, UK, Europe, China, Australia, Singapore | USA, Canada, UK, Europe, China, South Korea, South Africa, Australia, Singapore, Thailand, Iran, Israel | USA, UK | USA, Canada, UK, Europe, Israel, Iran, China, Taiwan, Australia |
| 6 | Most common predictors | Age, ethnicity, BMI, family history of diabetes, and prior GDM | - | Age, BMI, family history of diabetes, and prior GDM | - |
| 7 | Predictors that improve model performance | Systolic BP | TG, PAPP-A, and lipocalin-2 | Ethnicity, Previous pregnancy complications, VFT, SFT, PCOS | HbA1c, hsCRP, TC, LDL-C, visfatin, beta-hCG, AFP, TG |

**Table 2.**
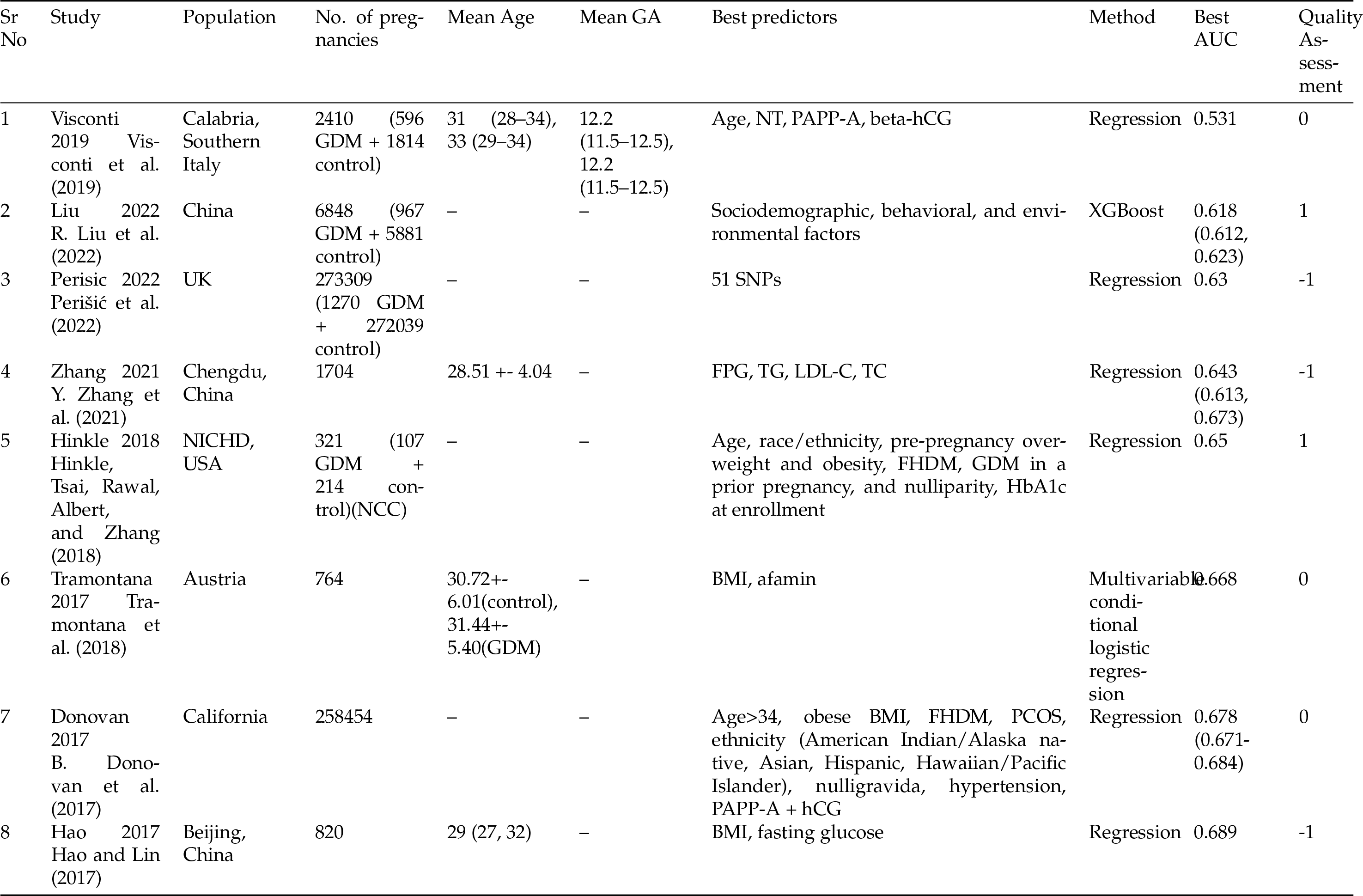
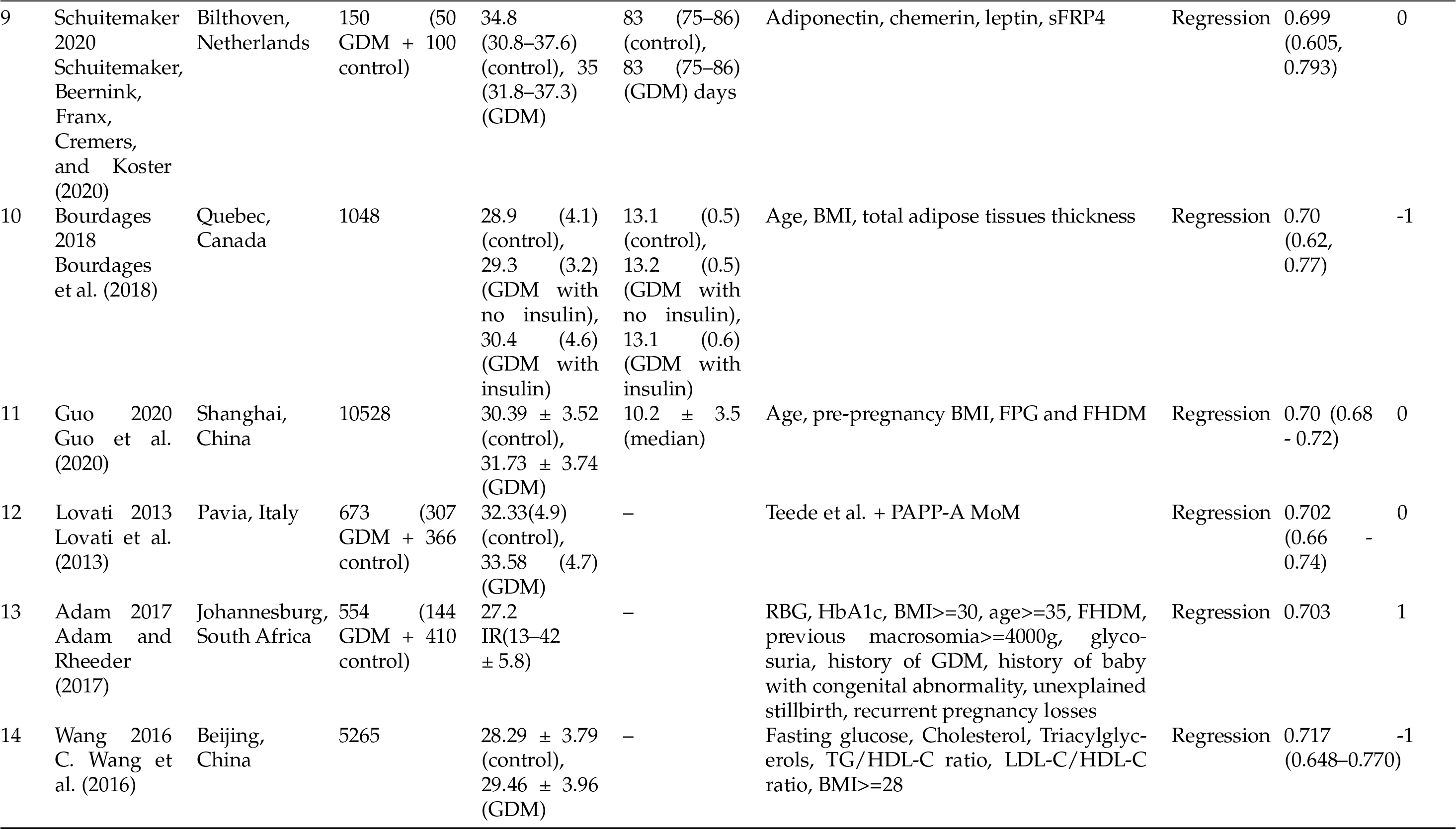
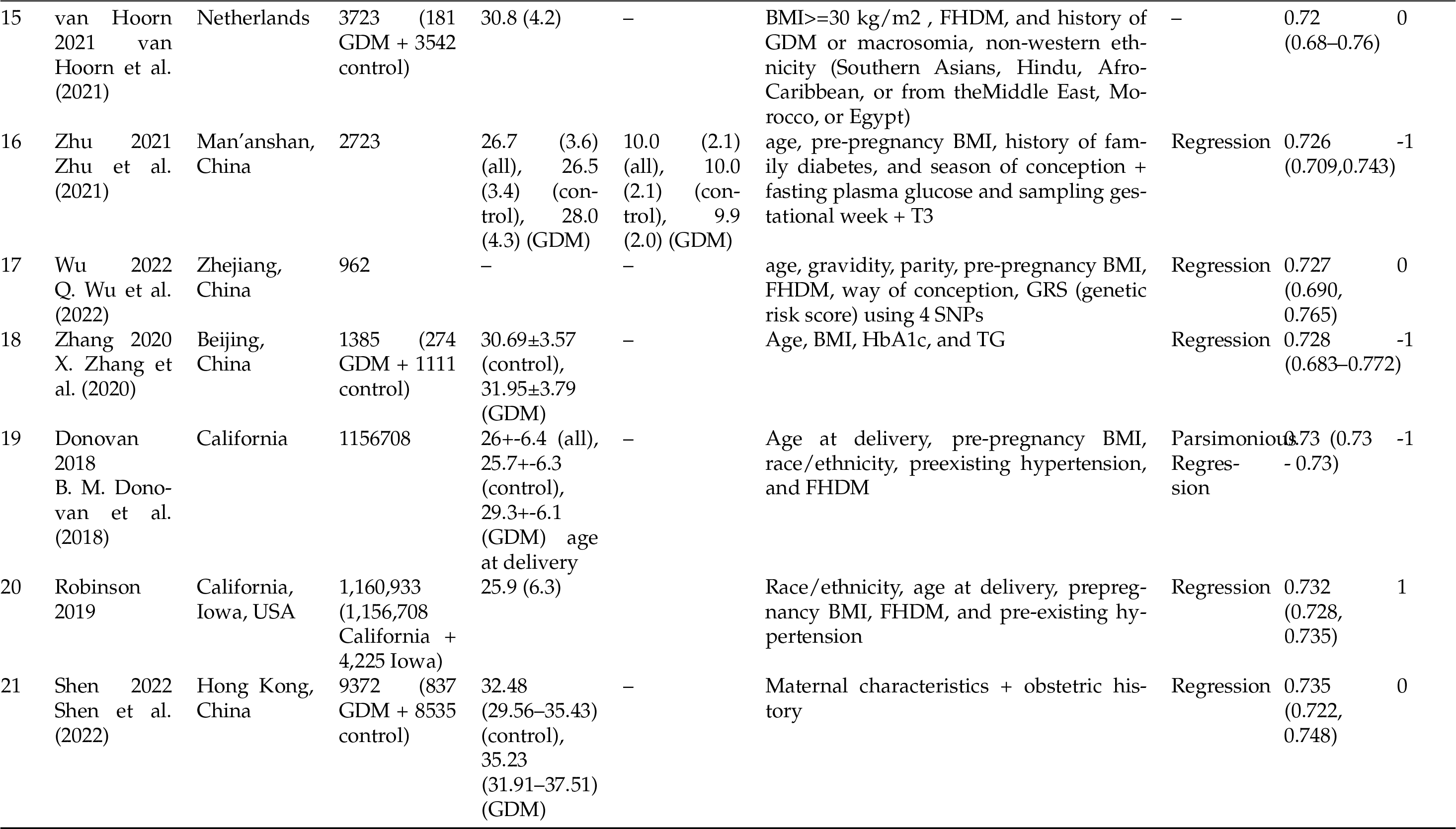
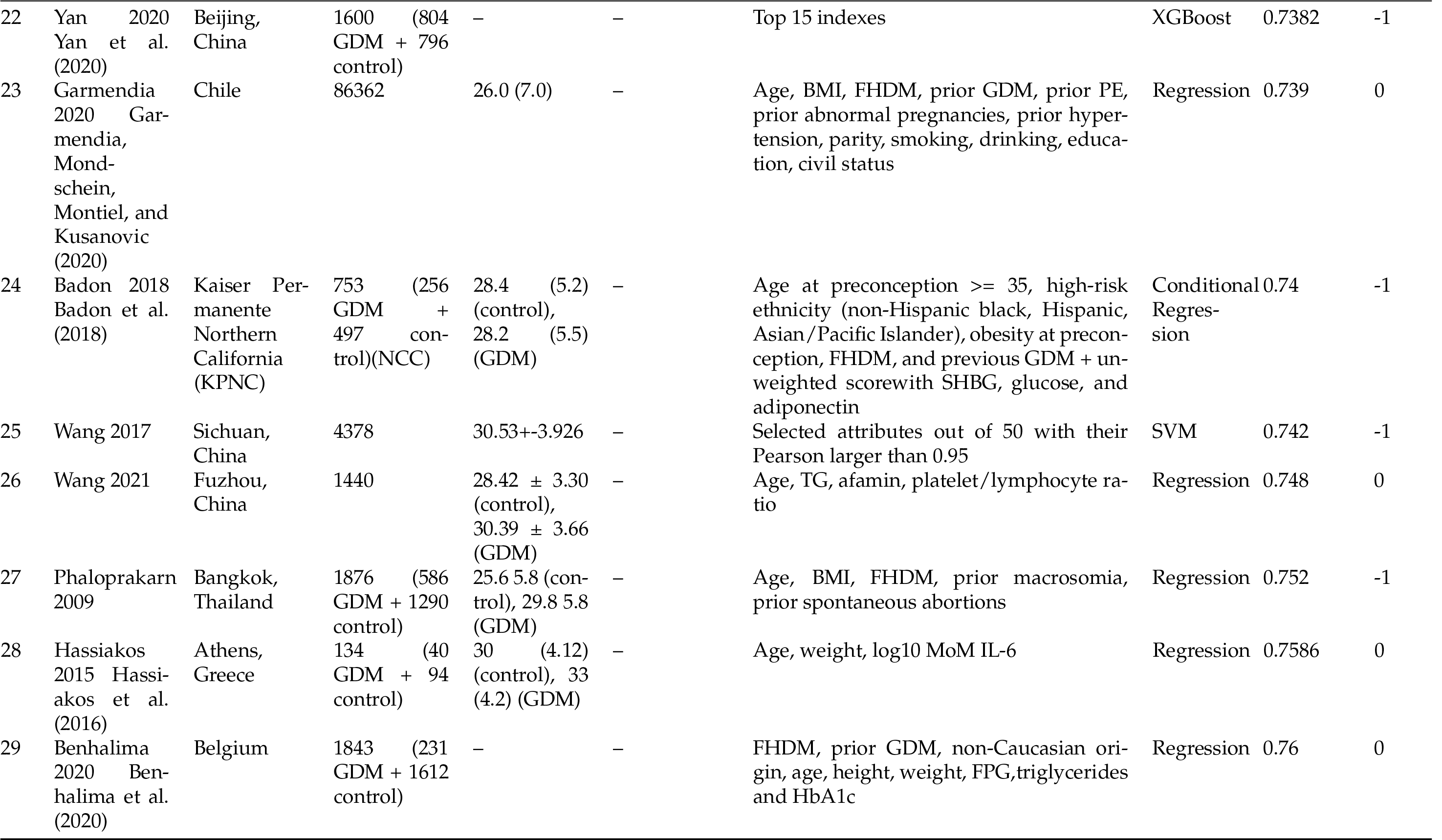
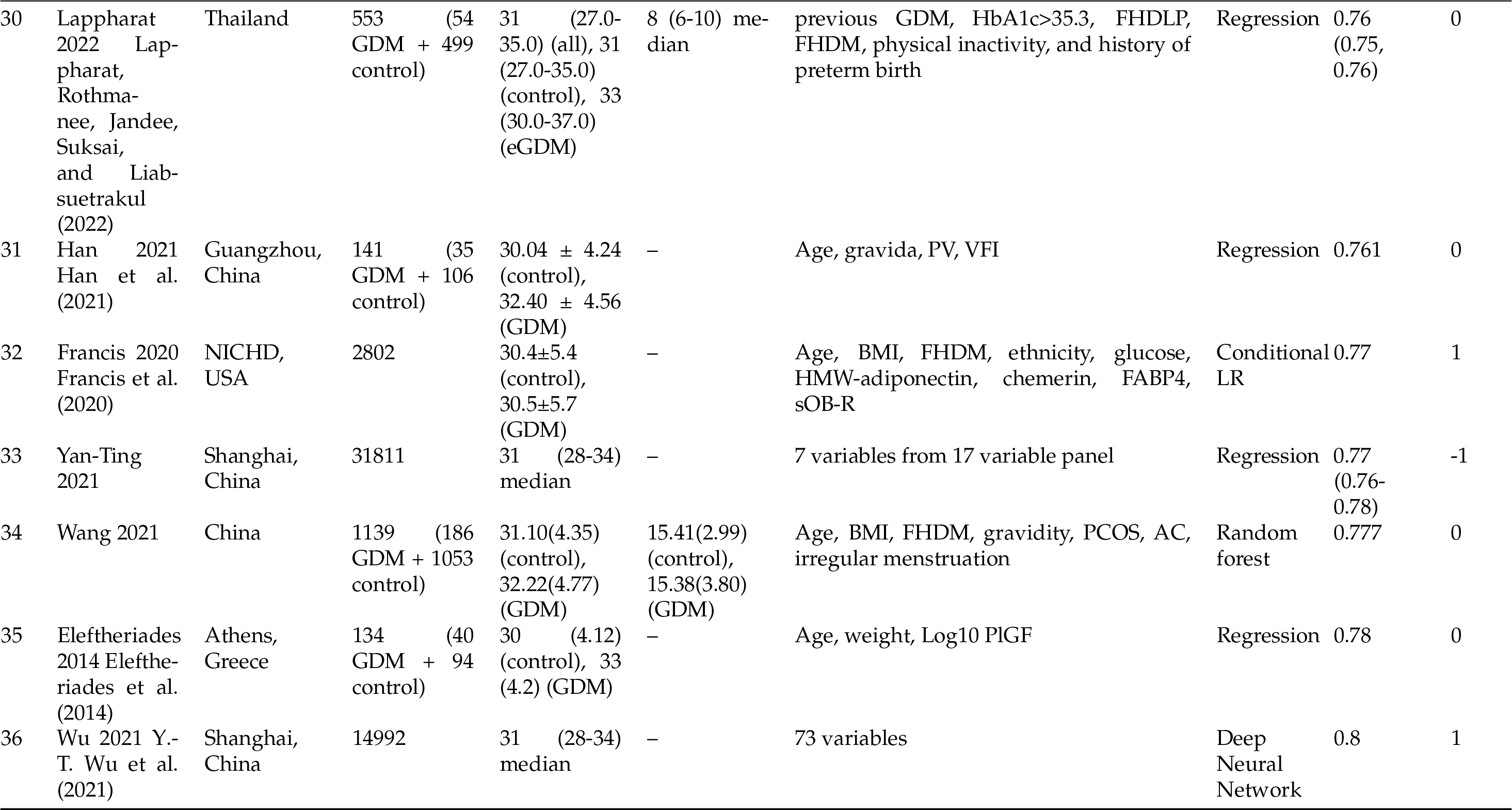
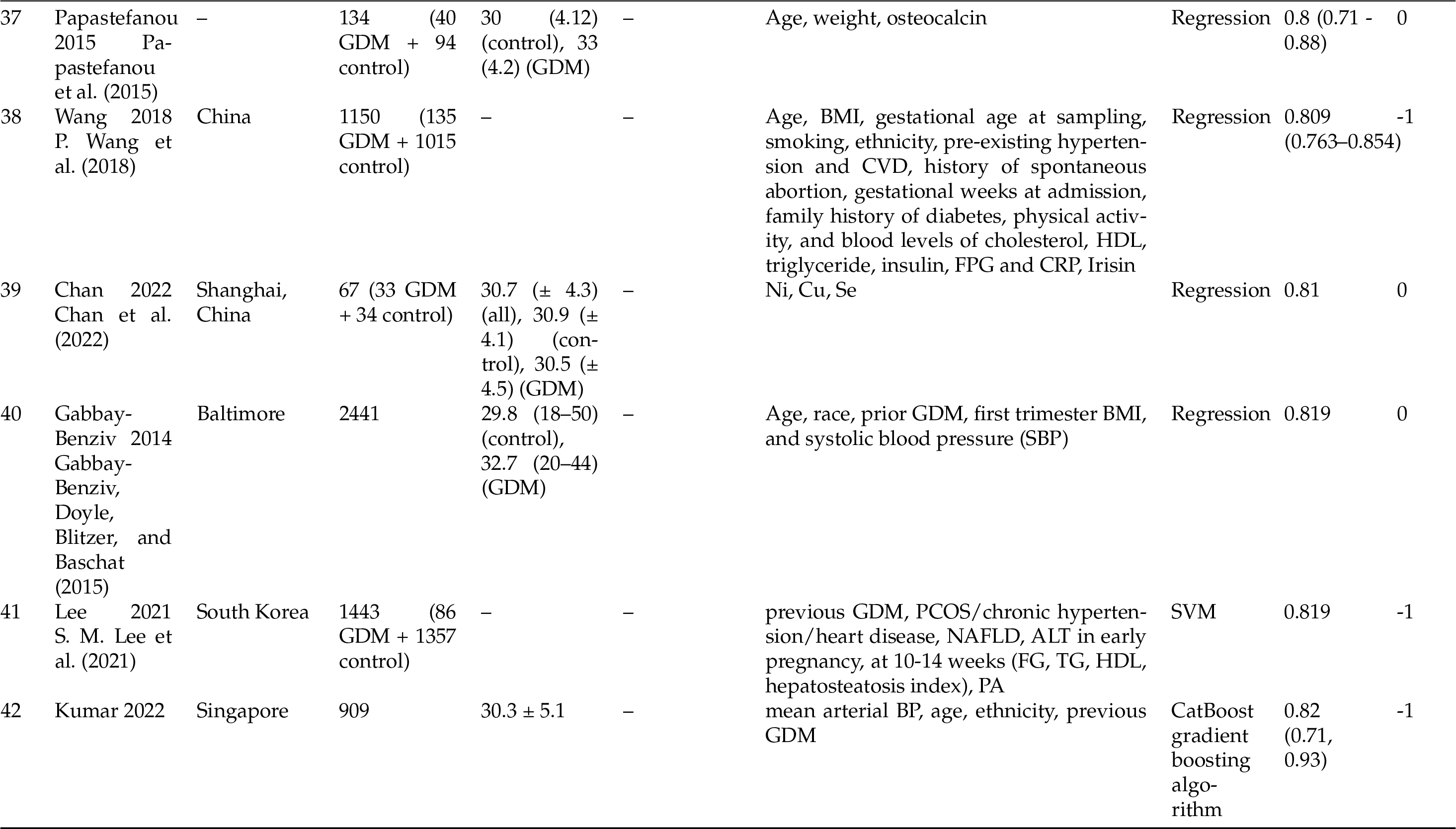
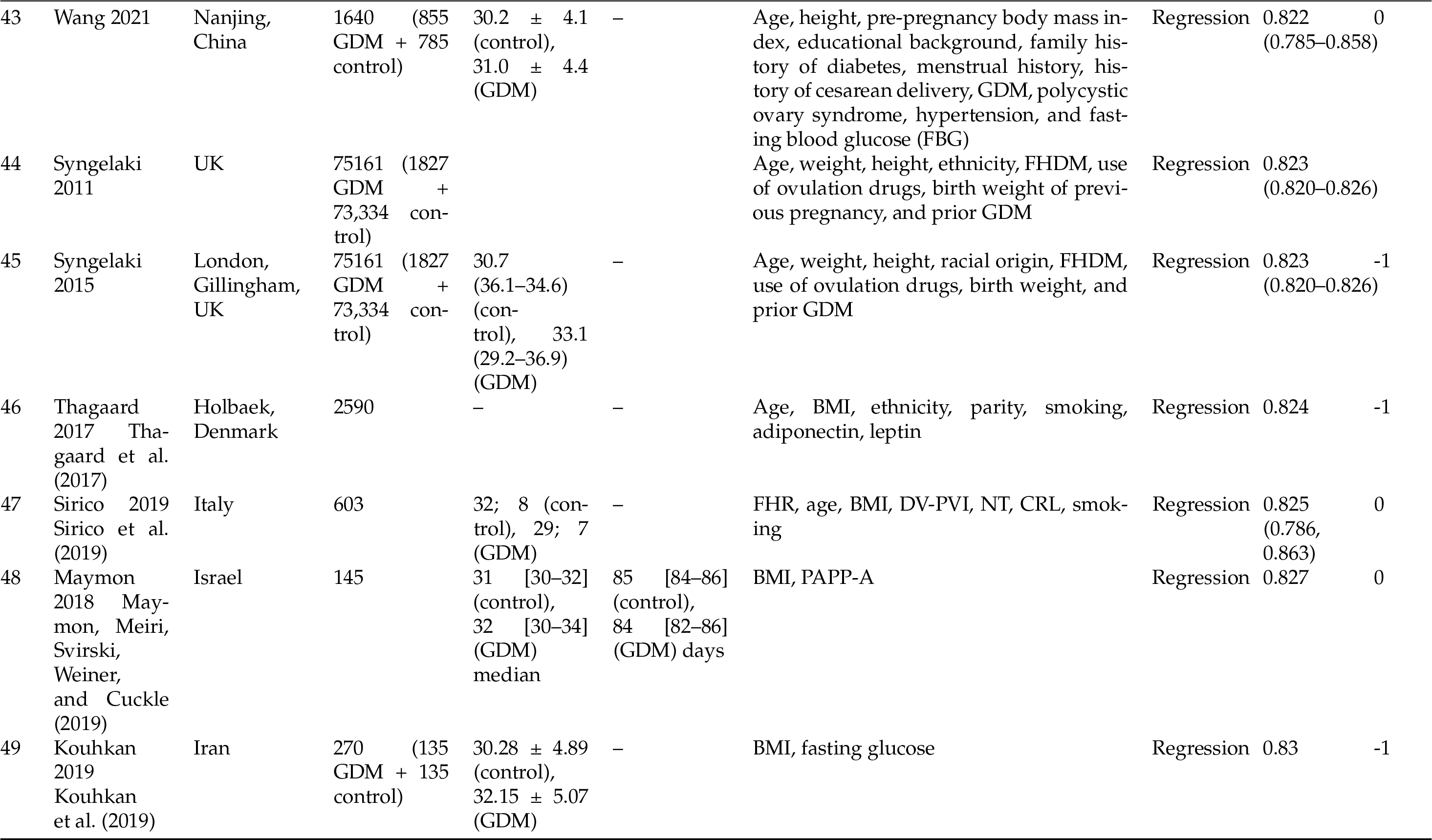
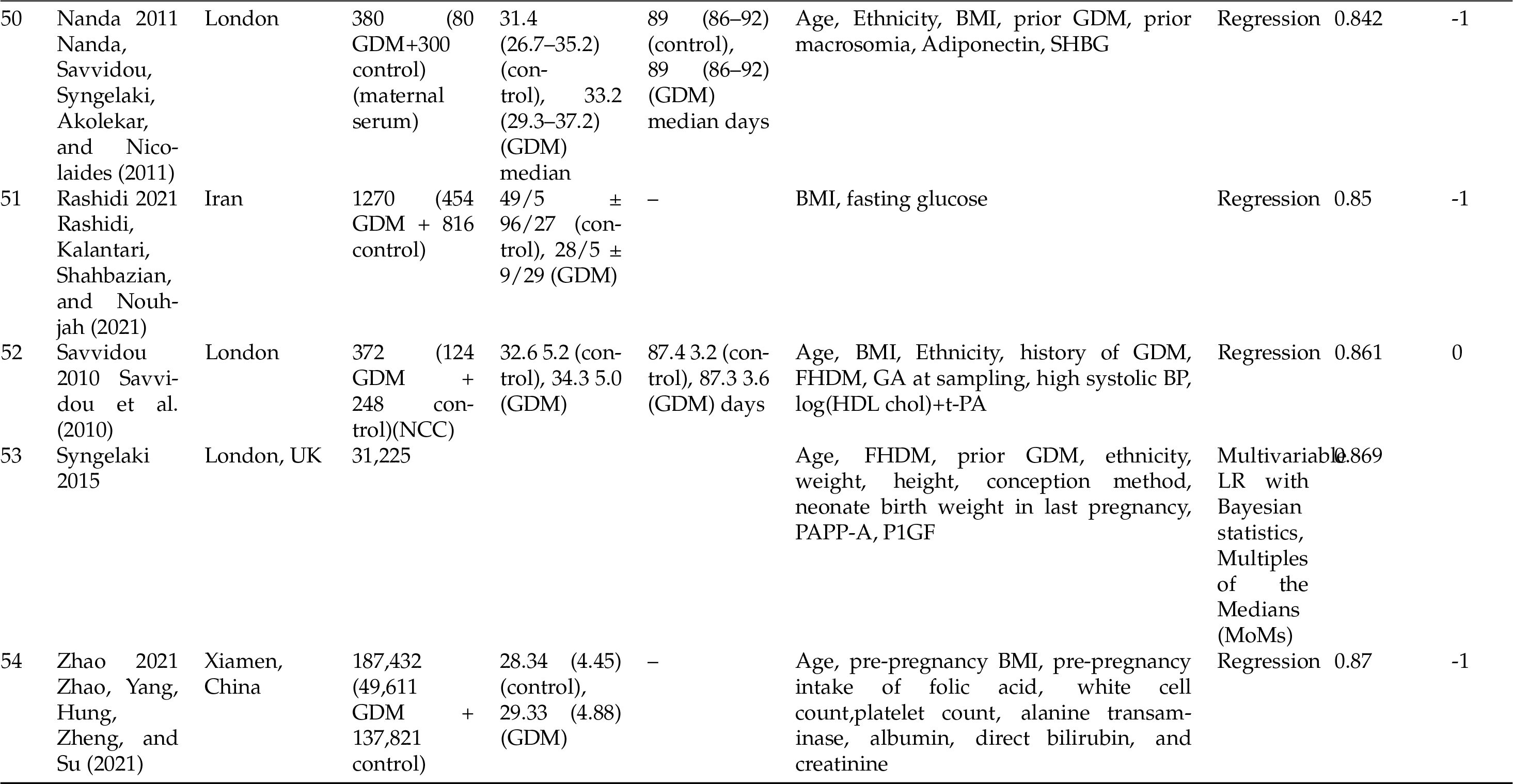
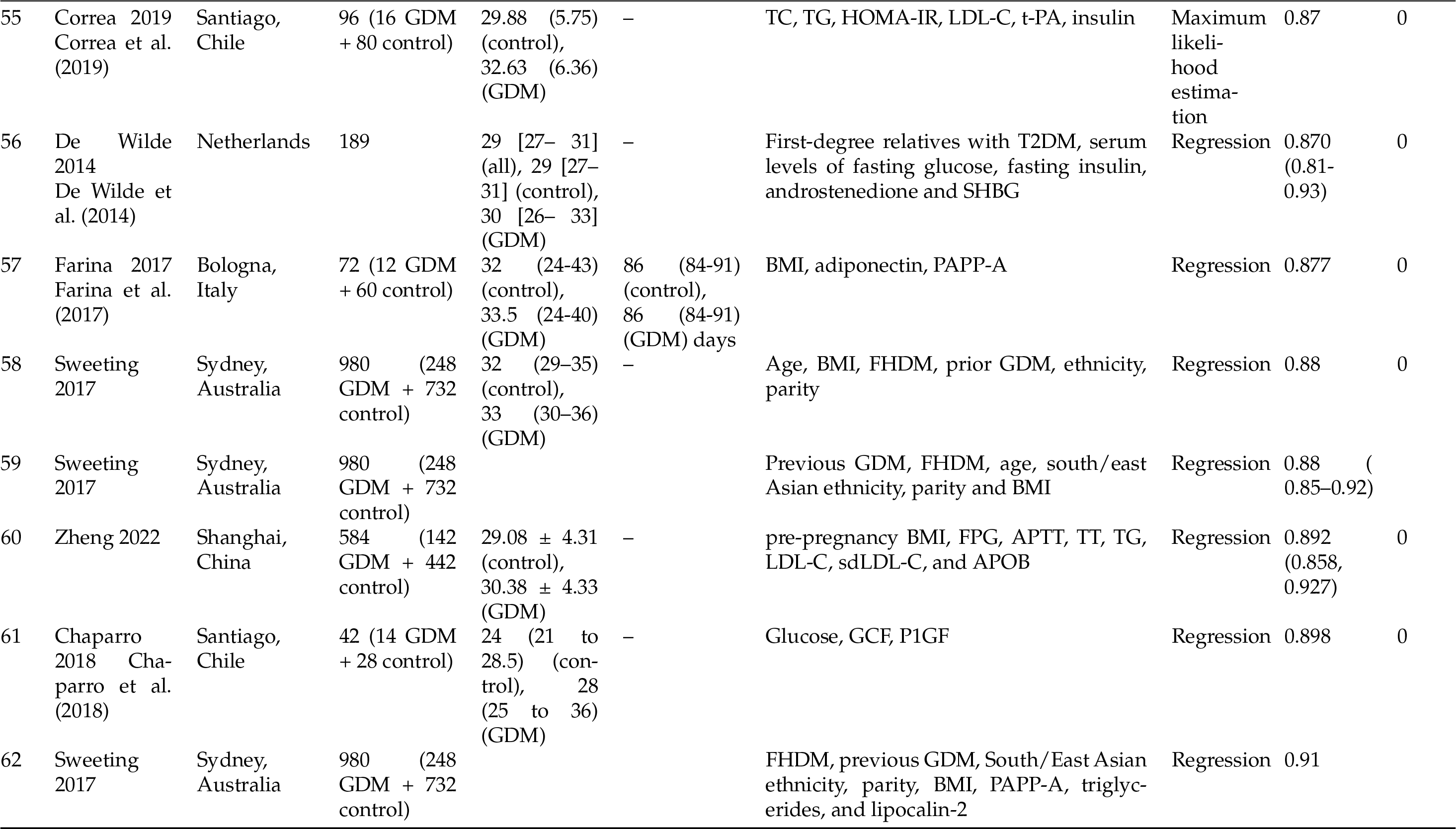
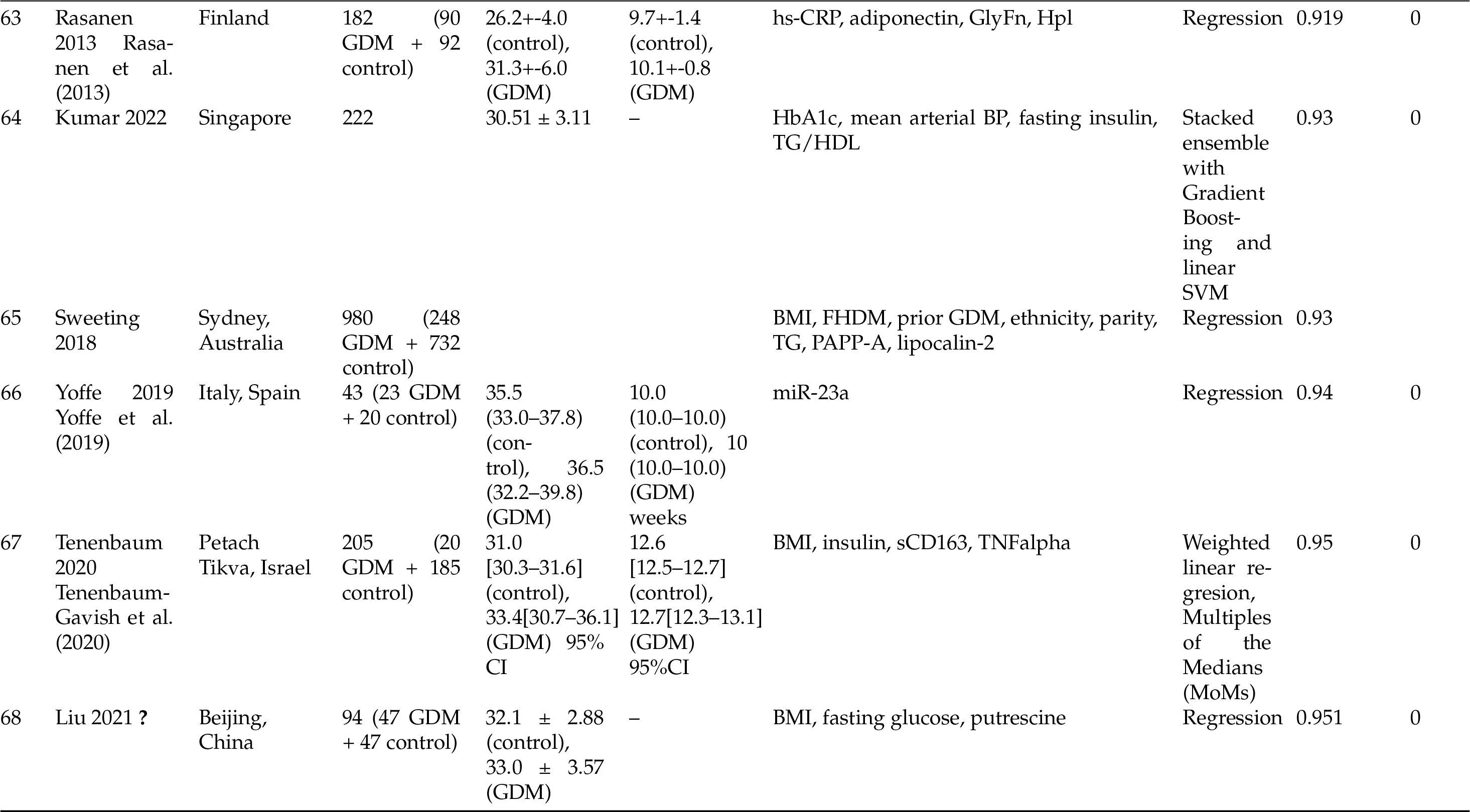

**Table 3.**
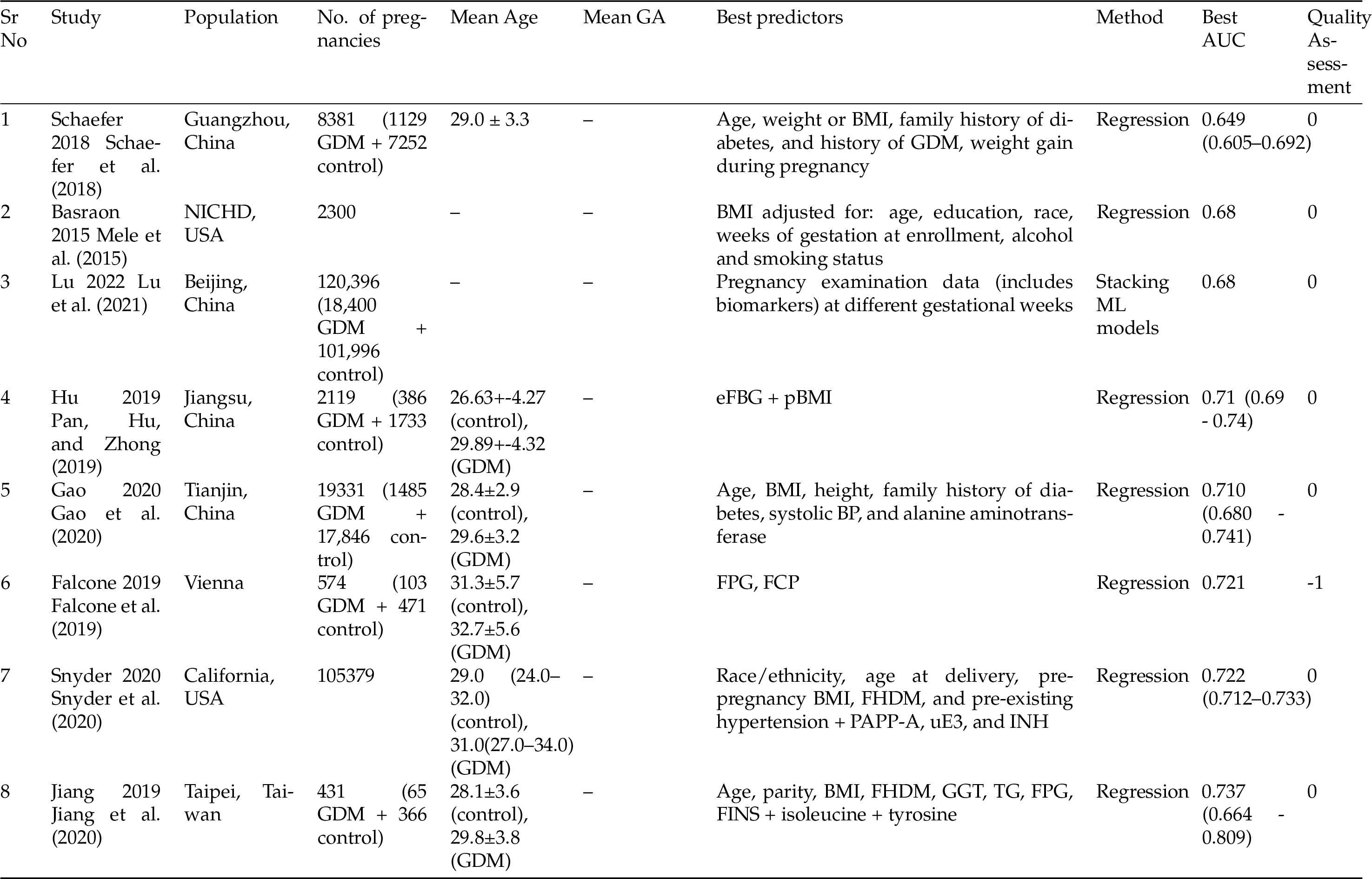
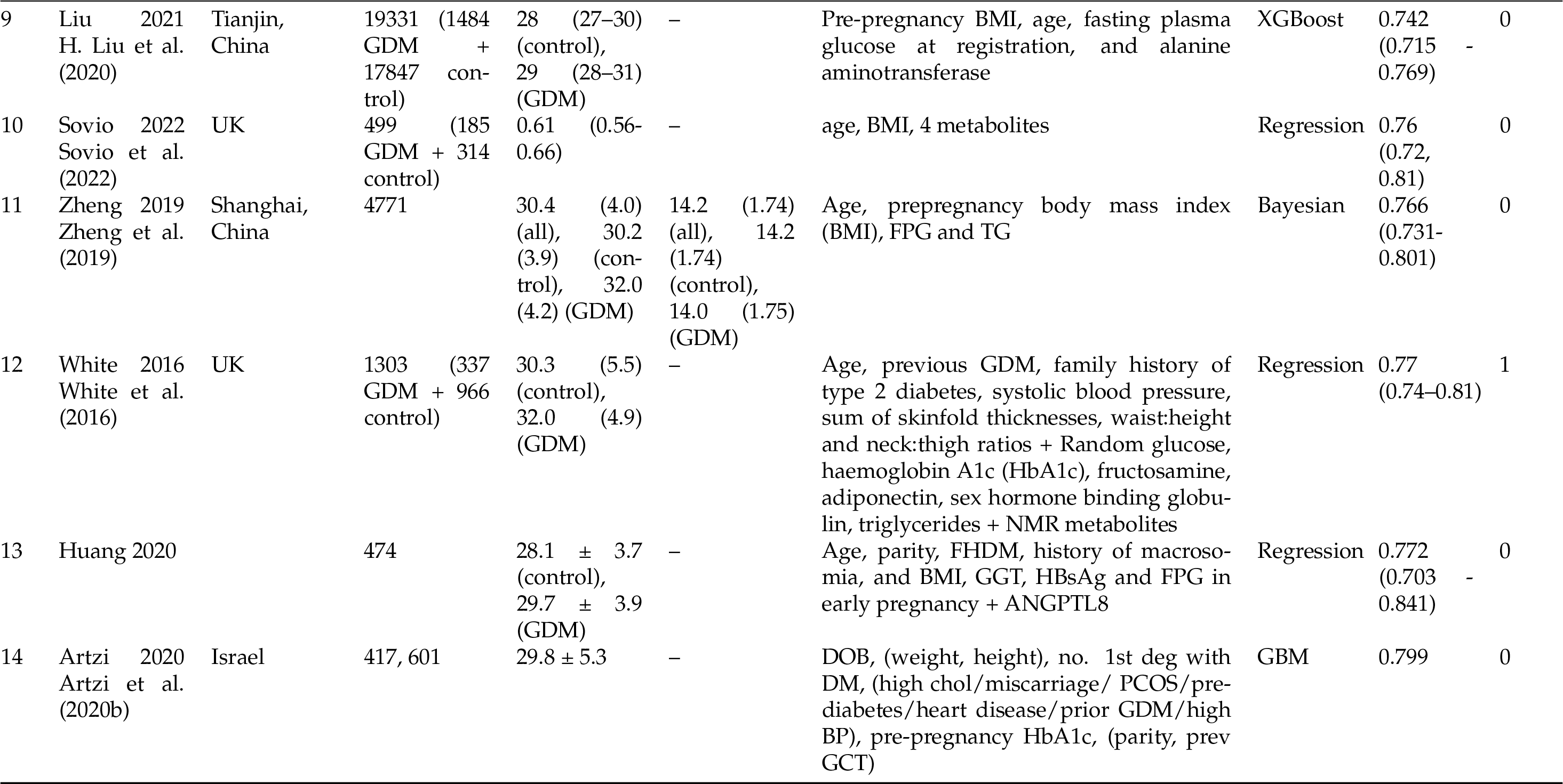
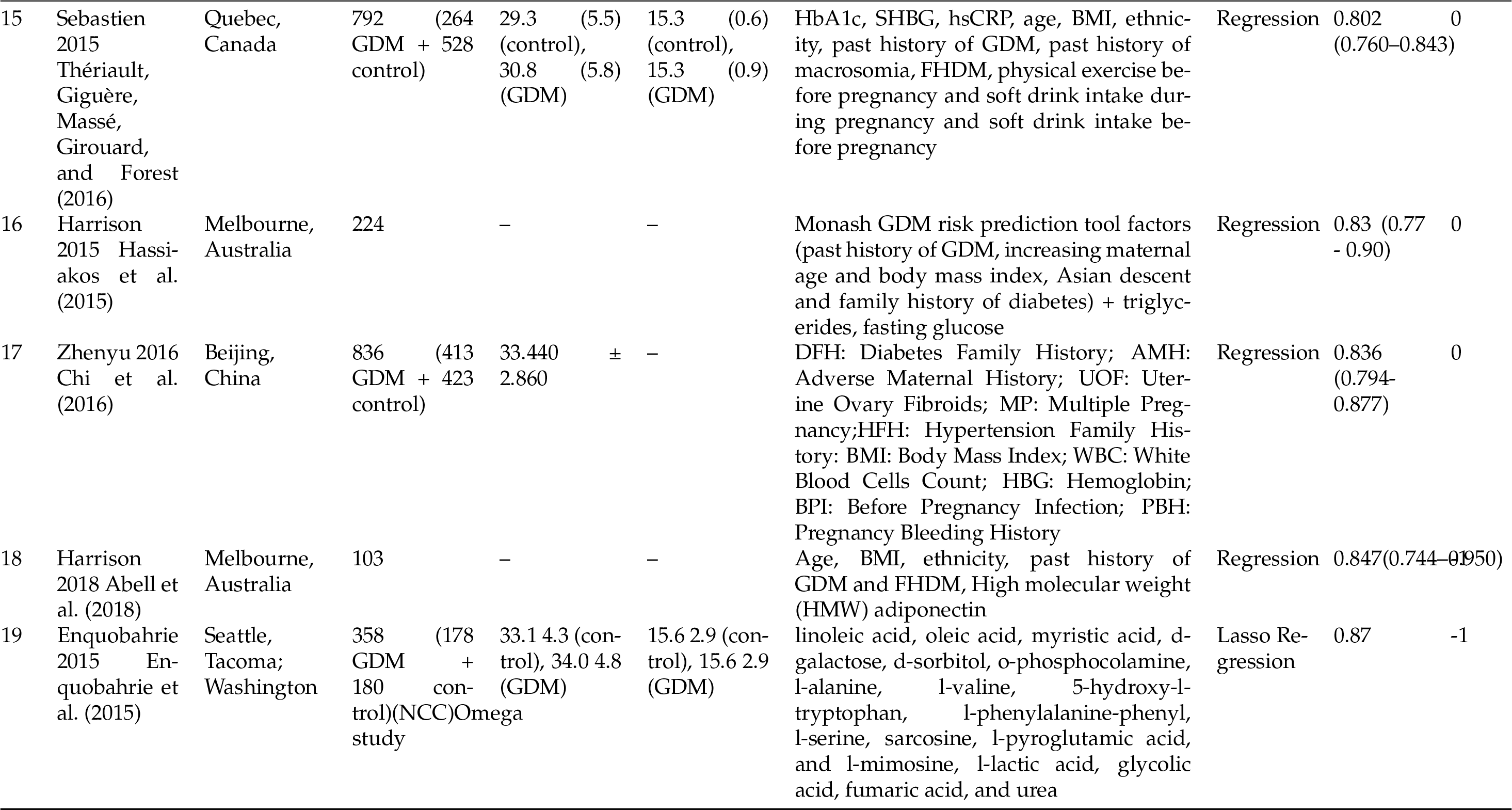
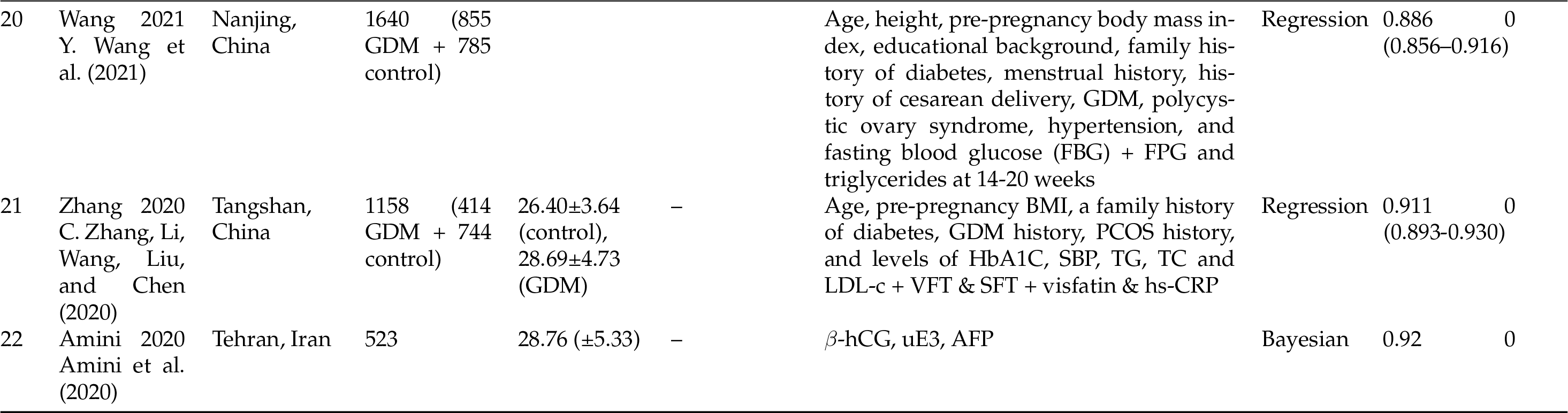

**Figure 1:**
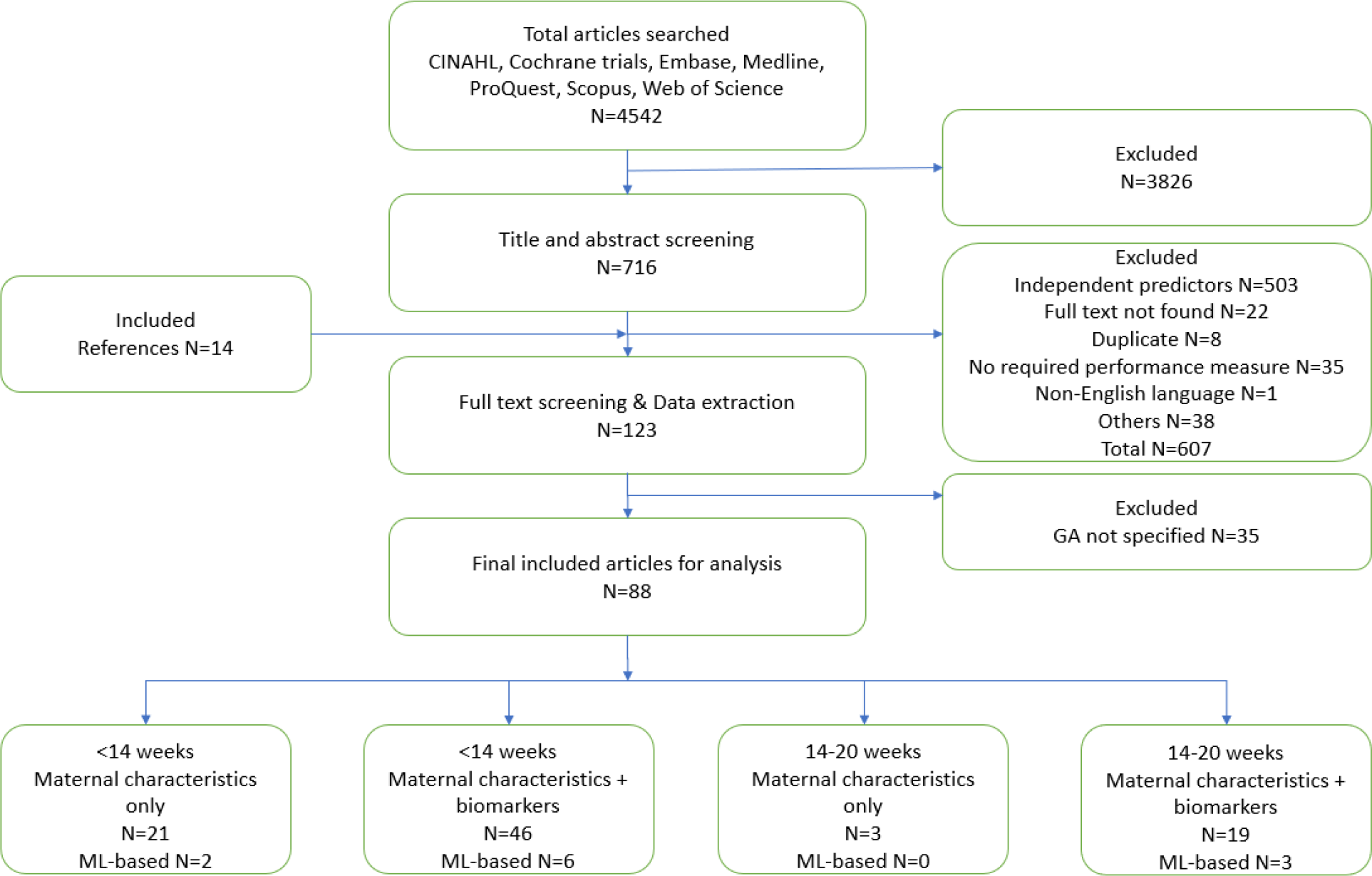
Consort Diagram.

The AUC also varied based on the population. age, ethnicity, BMI, family history of diabetes, and prior GDM were the most common maternal characteristics used. The addition of systolic blood pressure (BP) seems to improve performance. Triglycerides, PAPP-A, and lipocalin-2, combined with the other common maternal characteristics, have the highest predictive performance at <14 weeks. Supplement table 1 show a simplified summary of the studies, most commonly used risk predictors and the population studied 1. Of the 89 studies, only 22 (24.72%) presented calibration measures. Internal validation was performed in 29 studies (32.58%) using random split or k-fold cross-validation and bootstrapping. Only 15 studies out of 89 (16.85%) performed external validation.

### Quality Assessment

Detailed qua;ity assessment scores are shown in supplementary table xx and Figure 2. Eighty-three out of 89 studies, had a low bias of participants and six had moderate risk due to lack of information on data sources and inclusion and exclusion criteria. All studies had a low bias of predictors. Moderate risk of bias were observed in eight studies and was high in three additional studies, on describing the outcomes (eg., definition of diagnostic criteria for GDM) A total of 52 (58.43%) studies had a high bias of analysis, of which 37 were due to use of univariate regression to select the significant predictors to be used in multivariate analysis. Additional 13 (14.61%) studies were classified as having high risk of bias due to sample size (<50 cases in the GDM category) and two (2.25%) more due to inappropriate handling of missing data. All studies were thought to have a low bias in applicability.

**Figure 2:**
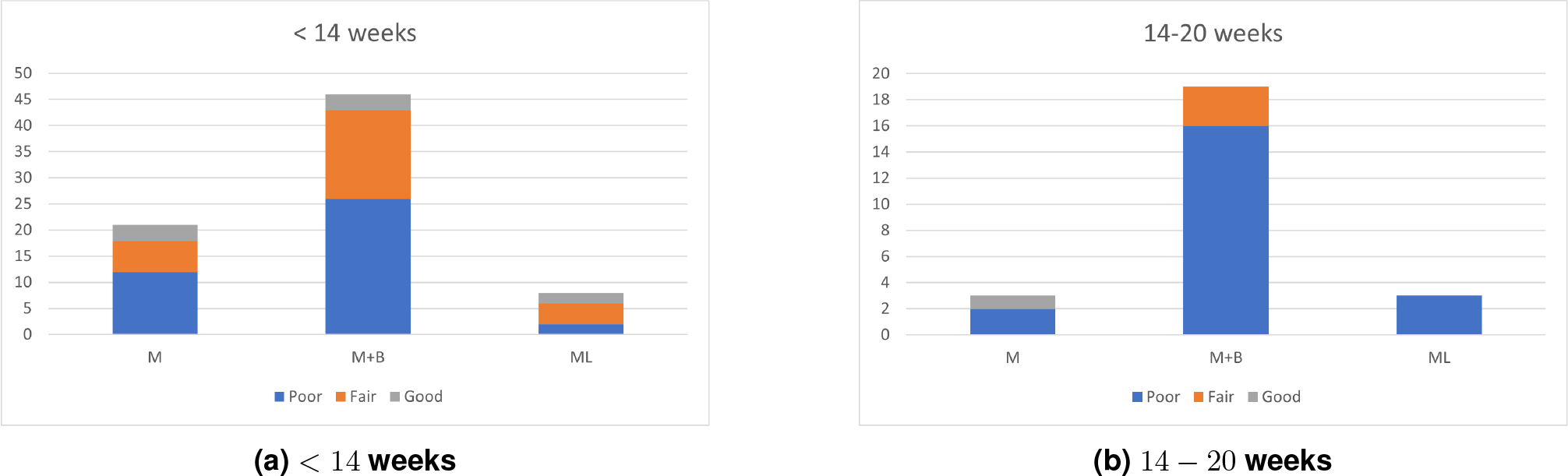
Quality Assessment of the included studies.

## Discussion

Our study is a comprehensive systematic review of all the published studies that used composite risk score prediction models for prediction of GDM in early gestation. Our key findings are: a) combining systolic BP with five commonly used risk factors such as age, ethnicity, BMI, family history of diabetes, and prior GDM improves the AUC for predicting GDM; b) inclusion of biomarkers with maternal characteristics improve the predictive performance; and c) use of ML-based techniques for prediction further improved the predictive scores, although the number of studies are limited.

Selective screening based on a composite risk scoring system is more effective in risk stratification of GDM women than a simple yes/no system based on standard risk factors. Risk factor-based approaches for screening for GDM are available, for example, Artzi et al. (2020a). However, it requires data from previous pregnancy, which might not always be practically available. Hence, we need a more practical approach to decide which women entering pregnancy will need testing for GDM.

Once such high-risk group is identified early on in pregnancy, clinicians can get sufficient time of 6-12 weeks to work towards prevention of GDM by implementing a proper diet and lifestyle intervention plan, as studied by Coomar et al. (2021). Overall, the performance of the prediction models published so far is not satisfactory to start implementing these in the clinics. The models purely based on maternal characteristics have low predictability and more research needs to be done to identify low-cost, yet highly predictive biomarkers. Moreover, very few studies used machine learning-based approach for building the prediction models Artzi et al. (2020a); Qiu et al. (2017); Y.-T. Wu et al. (2021); **?**); **?**, and this needs to be studied in greater detail, especially in the first trimester or at the time of first pregnancy visit, to unleash the existing predictive potential of the data and pave way for prevention strategies. Also, notable differences in the prediction performance of the models for populations of different origins justify the need for a population-based approach for developing early pregnancy prediction models for GDM.

The key strengths of our review are an exhaustive search strategy using a variety of databases & reference lists, and subgroup analysis based on time of collection of predictors - < 14 weeks or 14-20 weeks & type of predictors - maternal or maternal + biomarkers. This review can help to guide future research in terms of the most predictive features for GDM prediction in the first trimester. Analysis of the population using which the prediction models were derived highlighted the regions of the world understudied to develop population-specific prediction models. Exclusion of studies which did not report auROC values is a limitation of this review, although a common performance measure was needed for uniformity and comparison between studies.

## Conclusion

Many composite risk score-based prediction models have been developed in the last decade for early pregnancy prediction of GDM. There is a lot of heterogeneity in the predictors used to build these models, time at which these predictors were collected, measures used to quantify performance of these models, and definitions used to label the GDM outcome. This is a first review to assess composite risk score prediction models in the first trimester for GDM prediction for different populations. All models do not clear the PROBAST quality assessment check, especially due to insufficient validation. This review describes the need of good-quality, population-specific prediction models, using AI-based approaches to predict GDM early, adopt proper diet and lifestyle interventions, and prevent adverse complications associated with GDM for the mother and the fetus.

## Data Availability

All data produced in the present study are available upon reasonable request to the authors.

## Declarations

Some journals require declarations to be submitted in a standardized format. Please check the Instructions for Authors of the journal to which you are submitting to see if you need to complete this section. If yes, your manuscript must contain the following sections under the heading ‘Declarations’:

- Funding
- Conflict of interest/Competing interests (check journal-specific guidelines for which heading to use)
- Ethics approval
- Consent to participate
- Consent for publication
- Availability of data and materials
- Code availability
- Authors’ contributions

If any of the sections are not relevant to your manuscript, please include the heading and write ‘Not applicable’ for that section.

Editorial Policies for:

Springer journals and proceedings: https://www.springer.com/gp/editorial-policies

Nature Portfolio journals: https://www.nature.com/nature-research/editorial-policies

*Scientific Reports*: https://www.nature.com/srep/journal-policies/editorial-policies

BMC journals: https://www.biomedcentral.com/getpublished/editorial-policies

## Acknowledgements

This work was supported by the funding from Warwick-Novo Nordisk international Doctoral Training Program (DP and SS) and partly by Medical Research Council, UK (MR/R020981/1) (PS and YW).

## Notes

### Competing Interest Statement

The authors have declared no competing interest.

### Funding Statement

PS and YW are partly funded by Medical Research Council, UK (MR/R020981/1); DP and SS are funded by Warwick-Novo Nordisk international Doctoral Training Program.

