## Appendix for "Systematic Review of risk score prediction models using maternal characteristics with and without biomarkers for the prediction of GDM"

**Table A1. Summary of international criteria for diagnosis of GDM**

| Guidelines | Glucose load for OGTT  g | Fasting  Mg/dl  (Mmol/l) | 1-hr  Mg/dl  (Mmol/l) | 2-hr  Mg/dl  (Mmol/l) | 3-hr  Mg/dl  (Mmol/l) | |
| --- | --- | --- | --- | --- | --- | --- |
| **O’Sullivan and Mahan** (Bhavadharini *et al.*, 2016) | 100 | 90  (5.0) | 165  (9.2) | 145  (8.1) | | 125  (6.9) |
| World Health Organization (**WHO**)  0 | 75 | 126  (7.0) | - | 140  (7.8) | | - |
| American Diabetes Association (**ADA**) (Care and Suppl, 2019)  One step strategy  Two step strategy  Screening (50g GCT followed by 100g)  Diagnostic (**NDDG**)  (Any two elevated values)  Diagnostic (**Carpenter-Coustan**)  (Any two elevated values)  American College of Obstetricians and Gynecologists (**ACOG**) | 75  50  100  100  Any one elevated value in the two-step strategy | 92  (5.1)  -  105  (5.8)  95  (5.3) | 180  (10.0)  130  (7.2)  190  (10.6)  180  (10.0) | 153  (8.5)  -  165  (9.2)  155  (8.6) | | -  -  145  (8.0)  140  (7.8) |
| International Association of Diabetes and Pregnancy Study Groups (**IADPSG**) (Reddi Rani and Begum, 2016) | 75 | 92  (5.1) | 180  (10.0) | 153  (8.5) | | - |
| National Institute of Health and Care Excellence (**NICE**) (Reddi Rani and Begum, 2016) | 75 | 120  (5.6) | - | 140  (7.8) | | - |
| Australasian Diabetes In Pregnancy Society (**ADIPS**) (Nankervis *et al.*, 2008) | 75 | 92  (5.1) | 180  (10) | 153  (8.5) | | - |
| Diabetes in Pregnancy Study group India (**DIPSI**) (Reddi Rani and Begum, 2016) | 75 | - | - | 140  (7.8) | | - |
| **Canadian Diabetes Association** (Reddi Rani and Begum, 2016) | 75 | 95  (5.3) | 191  (10.6) | 160  (8.9) | | - |

**Table A2. Search terms**

| Database | Search strategy |
| --- | --- |
| Web of Science | ("gestational diabetes") OR (glucose intolerance AND pregnan*) OR (hyperglyc* NEAR/3 pregnan*) OR (diabetes NEAR/3 pregnan*)  AND  ("prediction model") OR ("statistic* model*") OR ("Machine learning") OR ("risk score") OR ("risk assessment tool*") |
| Scopus | (TITLE-ABS-KEY (("gestational diabetes") OR (glucose intolerance AND pregnan*) OR (hyperglyc* NEAR/3 pregnan*) OR (diabetes NEAR/3 pregnan*))  AND  TITLE-ABS-KEY (("prediction model”) OR ( "statistic* model*" )  OR  ( "Machine learning" )  OR  ( "risk score" )  OR  ( "risk assessment tool*" ) ) ) |
| CINAHL | TX ( ("gestational diabetes") OR (glucose intolerance AND pregnan*) OR (hyperglyc* NEAR/3 pregnan*) OR (diabetes NEAR/3 pregnan*) )  AND  TX ( ("prediction model") OR ("statistic* model*") OR ("Machine learning") OR ("risk score") OR ("risk assessment tool*") ) |
| Cochrane | ("gestational diabetes") OR (glucose intolerance AND pregnan*) OR (hyperglyc* NEAR/3 pregnan*) OR (diabetes NEAR/3 pregnan*) in Title Abstract Keyword  AND  ("prediction model") OR ("statistic* model*") OR ("Machine learning") OR ("risk score") OR ("risk assessment tool*") in Title Abstract Keyword - (Word variations have been searched) |
| ProQuest | ("gestational diabetes") OR (glucose intolerance AND pregnan*) OR (hyperglyc* NEAR/3 pregnan*) OR (diabetes NEAR/3 pregnan*)  AND  ("prediction model") OR ("statistic* model*") OR ("Machine learning") OR ("risk score") OR ("risk assessment tool*")  Databases searched: SciTech Premium Collection, Biological Science Collection, Natural Science Collection, Biological Science Database, Science Database, Biological Science Index |
| Ethos | (gestational diabetes) AND (prediction) |
| OpenGrey | (gestational diabetes) AND (prediction) |

Figure A1. Search strategy

a. Medline:

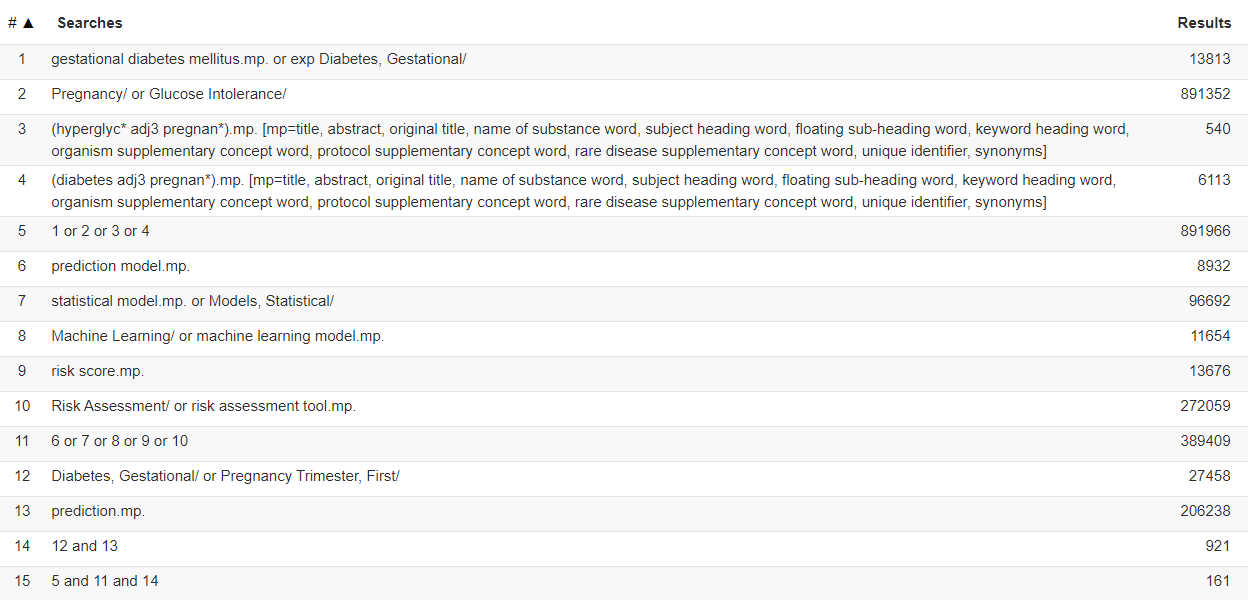

b. Embase:

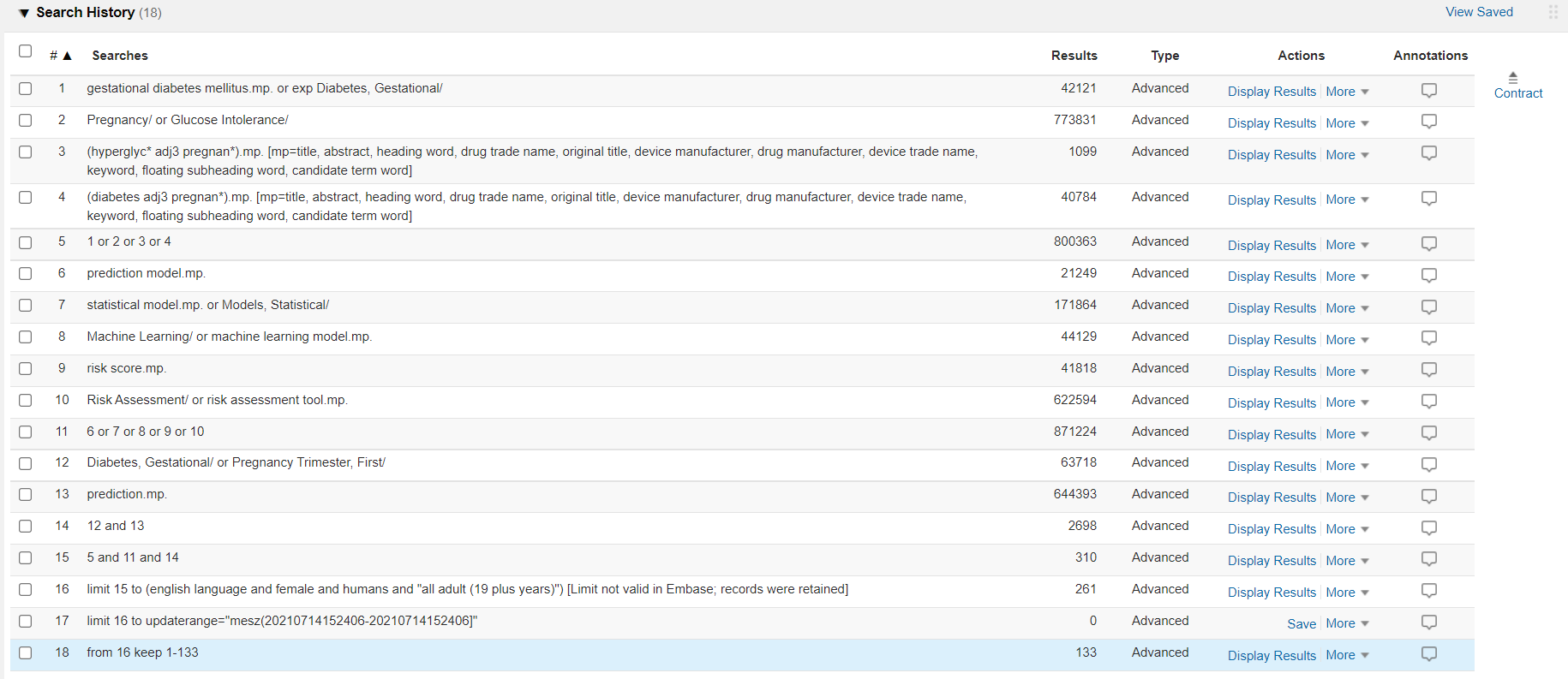

**Table A4. Selection criteria of predictive modelling studies in PICOTS format**

a. <14 weeks

|  | **Participants (P)** | **Intervention**  **(I)** | **Control (C)** | **Outcomes**  **(O)** | **Timeframe**  **(T)** | **Setting**  **(S)** | **Other limits** |
| --- | --- | --- | --- | --- | --- | --- | --- |
| **Inclusion criteria** | Pregnant women or planning to get pregnant | Composite risk score prediction models | Gold standard methods for GDM diagnosis (Table A1) | **Primary:** metrics of discrimination ability (auROC, sensitivity, specificity, ppv, and npv), calibration, and classification accuracy in GDM prediction  **Secondary:** important variables, intended use of models, risk equations and their cut-offs | All studies till July 2021 | **Clinical care settings** e.g. hospitals, institutions, case-control study, population-based cohort. | English language |
| **Exclusion criteria** | Patients with other clinical phenotypes of diabetes; Diabetic complication. |  |  | Papers with no mention of auROC. |  |  | Other language |

**Table A5. Risk of bias and applicability assessment by PROBAST criteria**

a. <14 weeks

| **Sr. no.** | **Study** | **Year** | **Type of prediction study** | **Type of trial** | **No. of  pregnancies** | **Participants** | | **Predictors** | | | **Outcome** | | | | | | | **Analysis** | | | | | | | | | | **Applicability** |
| --- | --- | --- | --- | --- | --- | --- | --- | --- | --- | --- | --- | --- | --- | --- | --- | --- | --- | --- | --- | --- | --- | --- | --- | --- | --- | --- | --- | --- |
|  |  |  |  |  |  | 1 | 2 | 1 | 2 | 3 | 1 | 2 | 3 | 4 | 5 | 6 | 1 | | 2 | 3 | 4 | 5 | 6 | 7 | 8 | 9 |  | |
| 1 | Hinkle | 2018 | Dev | NCC | 321 | 1 | 1 | 1 | 1 | 1 | 1 | 1 | 1 | 1 | 1 | 1 | 1 | | 1 | 0 | 1 | 1 | 1 | 1 | -1 | 1 | 1 | |
| 2 | Adam | 2017 | Dev | Cohort | 554 | 1 | 1 | 1 | 1 | 1 | 1 | 1 | 1 | 1 | 1 | 1 | 1 | | 1 | 1 | 1 | 1 | 1 | 1 | -1 | 1 | 1 | |
| 3 | Yan | 2020 | Dev |  | 1600 | -1 | -1 | 1 | 1 | 1 | -1 | -1 | 1 | 1 | 1 | 1 | 1 | | 1 | 0 | -1 | 1 | 1 | 1 | 1 | 1 | 1 | |
| 4 | Badon | 2018 | Dev | NCC | 753 | 1 | 1 | 1 | 1 | 1 | 1 | 1 | 1 | 1 | 1 | 1 | 1 | | 1 | 0 | -1 | 1 | 1 | 1 | -1 | 1 | 1 | |
| 5 | Wang | 2017 | Dev | NCC | 4378 | 1 | 1 | 1 | 1 | 1 | 1 | 1 | 1 | 1 | 1 | 1 | 1 | | -1 | 0 | 1 | 1 | 1 | 1 | 1 | 1 | 1 | |
| 6 | Hassiakos | 2015 | Dev | Case-control | 134 | 1 | 1 | 1 | 1 | 1 | 1 | 1 | 1 | 1 | 1 | 1 | 1 | | 1 | 1 | -1 | 1 | 1 | 1 | -1 | 1 | 1 | |
| 7 | Benhalima | 2020 | Dev | Prospective cohort | 1843 | 1 | 1 | 1 | 1 | 1 | 1 | 1 | 1 | 1 | 1 | 1 | 1 | | 1 | 1 | 1 | 0 | 1 | 1 | -1 | 1 | 1 | |
| 8 | Eleftheriades | 2014 | Dev | Case-control | 134 | 1 | 1 | 1 | 1 | 1 | 1 | 1 | 1 | 1 | 1 | 1 | 1 | | 1 | 1 | -1 | 1 | 1 | 1 | -1 | 1 | 1 | |
| 9 | Gabbay- Benziv | 2014 | Dev | Prospective cohort | 2441 | 1 | 1 | 1 | 1 | 1 | 1 | 1 | 1 | 1 | 1 | 1 | 0 | | 1 | 0 | 1 | 0 | 1 | 1 | -1 | 1 | 1 | |
| 10 | Lee | 2021 | Dev | Prospective cohort | 1443 | 1 | 1 | 1 | 1 | 1 | -1 | -1 | 1 | 1 | 1 | 1 | 0 | | -1 | 1 | -1 | 1 | 1 | 1 | -1 | 1 | 1 | |
| 11 | Nanda | 2011 | Dev | Case-control | 380 | 1 | 1 | 1 | 1 | 1 | 1 | 1 | 1 | 1 | 1 | 1 | 1 | | 1 | 0 | -1 | 1 | 1 | 1 | -1 | 1 | 1 | |
| 12 | Savvidou | 2010 | Dev | NCC | 372 | 1 | 1 | 1 | 1 | 1 | 1 | 1 | 1 | 1 | 1 | 1 | 1 | | 1 | 1 | 1 | 0 | 1 | 1 | -1 | 1 | 1 | |
| 13 | Zhao | 2021 | Dev | Cohort | 187432 | 1 | 1 | 1 | 1 | 1 | -1 | 1 | 1 | 1 | 1 | 1 | 1 | | 1 | 1 | 1 | 1 | 1 | 1 | -1 | 1 | 1 | |
| 14 | Sweeting | 2017 | Dev | Retrospective case-control | 980 | 1 | 1 | 1 | 1 | 1 | 1 | 1 | 1 | 1 | 1 | 1 | 1 | | 1 | 1 | 1 | 0 | 1 | 1 | -1 | 1 | 1 | |
| 15 | Yoffe | 2019 | Dev & Val | Case-control | 43 | 1 | 1 | 1 | 1 | 1 | 1 | 1 | 1 | 1 | 1 | 1 | 1 | | 1 | 1 | -1 | 1 | 1 | 1 | -1 | 1 | 1 | |
| 16 | Donovan | 2017 | Dev | Cohort | 258454 | 1 | 1 | 1 | 1 | 1 | 1 | -1 | 1 | 1 | 1 | 1 | 1 | | -1 | 1 | -1 | 0 | 1 | 1 | -1 | 1 | 1 | |
| 17 | Guo | 2020 | Dev | Prospective cohort | 10528 | 1 | 1 | 1 | 1 | 1 | 1 | 1 | 1 | 1 | 1 | 1 | 1 | | 1 | 0 | 1 | 0 | 1 | 1 | -1 | 1 | 1 | |
| 18 | Lovati | 2013 | Dev | Case-control | 673 | 1 | 1 | 1 | 1 | 1 | 1 | 1 | 1 | 1 | 1 | 1 | 1 | | 1 | 1 | -1 | 0 | 1 | 1 | -1 | 1 | 1 | |
| 19 | Wang | 2016 | Dev | Retrospective | 5265 | 1 | 1 | 1 | 1 | 1 | 1 | 1 | 1 | 1 | 1 | 1 | 1 | | 1 | 1 | 1 | 1 | 1 | 1 | -1 | 1 | 1 | |
| 20 | van Hoorn | 2021 | Dev | Prospective multicentre cohort | 3723 | 1 | 1 | 1 | 1 | 1 | 1 | 1 | 1 | 1 | 1 | 1 | 0 | | -1 | 1 | -1 | 0 | 1 | 1 | -1 | 0 | 1 | |
| 21 | Zhu | 2021 | Dev | Prospective cohort | 2723 | 1 | 1 | 1 | 1 | 1 | 1 | 1 | 1 | 1 | 1 | 1 | 1 | | 1 | 1 | -1 | 1 | 1 | 1 | -1 | 1 | 1 | |
| 22 | Zhang | 2020 | Dev | Prospective cohort | 1385 | 1 | 1 | 1 | 1 | 1 | 1 | 1 | 1 | 1 | 1 | 1 | 1 | | -1 | 1 | -1 | 1 | 1 | 1 | -1 | 1 | 1 | |
| 23 | Donovan | 2018 | Dev | Cohort | 1156708 | 1 | 1 | 1 | 1 | 1 | -1 | -1 | 1 | 1 | 1 | 1 | -1 | | -1 | 1 | -1 | 1 | 1 | 1 | -1 | 1 | 1 | |
| 24 | Robinson | 2019 | Dev & Val | Cohort | 11,60,933 | 1 | 1 | 1 | 1 | 1 | 1 | 1 | 1 | 1 | 1 | 1 | 0 | | 1 | 0 | 1 | 1 | 1 | 1 | -1 | 1 | 1 | |
| 25 | Yan-Ting | 2021 | Dev |  | 31811 | 1 | 1 | 1 | 1 | 1 | 1 | 1 | 1 | 1 | 1 | 1 | 1 | | -1 | 1 | -1 | 1 | 1 | 1 | 1 | 1 | 1 | |
| 26 | Papastefanou | 2015 | Dev | Prospective case-control | 134 | 1 | 1 | 1 | 1 | 1 | 1 | 1 | 1 | 1 | 1 | 1 | 1 | | 1 | 1 | -1 | 1 | 1 | 1 | -1 | 1 | 1 | |
| 27 | Wang | 2018 | Dev | Prospective multicentre cohort | 1150 | 1 | 1 | 1 | 1 | 1 | 1 | 1 | 1 | 1 | 1 | 1 | 1 | | 1 | 1 | -1 | 1 | 1 | 1 | -1 | 1 | 1 | |
| 28 | Wang | 2021 | Dev | Retrospective cohort | 1640 | 1 | 1 | 1 | 1 | 1 | 1 | 1 | 1 | 1 | 1 | 1 | 1 | | 1 | 1 | -1 | 0 | 1 | 1 | -1 | 1 | 1 | |
| 29 | Syngelaki | 2015 | Dev | Prospective screening | 75161 | 1 | 1 | 1 | 1 | 1 | 1 | -1 | 1 | 1 | 1 | 1 | 1 | | 1 | 1 | -1 | 1 | 1 | 1 | -1 | 1 | 1 | |
| 30 | De Wilde | 2014 | Dev | Prospective multicentre cohort | 189 | 1 | 1 | 1 | 1 | 1 | 1 | 1 | 1 | 1 | 1 | 1 | 1 | | 1 | 0 | 1 | 0 | 1 | 1 | -1 | 1 | 1 | |
| 31 | Sweeting | 2017 | Dev | Retrospective case-control | 980 | 1 | 1 | 1 | 1 | 1 | 1 | 1 | 1 | 1 | 1 | 1 | 1 | | 1 | 1 | 1 | 0 | 1 | 1 | -1 | 1 | 1 | |

b. 14-20 weeks

| **Sr. no.** | **Study** | **Year** | **Type of prediction study** | **Type of trial** | **No. of  pregnancies** | **Participants** | | **Predictors** | | | **Outcome** | | | | | | **Analysis** | | | | | | | | | **Applicability** |
| --- | --- | --- | --- | --- | --- | --- | --- | --- | --- | --- | --- | --- | --- | --- | --- | --- | --- | --- | --- | --- | --- | --- | --- | --- | --- | --- |
|  |  |  |  |  |  | 1 | 2 | 1 | 2 | 3 | 1 | 2 | 3 | 4 | 5 | 6 | 1 | 2 | 3 | 4 | 5 | 6 | 7 | 8 | 9 |  |
| 1 | Basraon | 2015 | Dev | Multicenter RCT | 2300 | 1 | 1 | 1 | 1 | 1 | 1 | 1 | 1 | 1 | 1 | 1 | 0 | 1 | 1 | -1 | 0 | 1 | 1 | -1 | 1 | 1 |
| 2 | Falcone | 2019 | Dev | Prospective cohort | 574 | 1 | 1 | 1 | 1 | 1 | 1 | 1 | 1 | 1 | 1 | 1 | 1 | 1 | 1 | -1 | 1 | 1 | 1 | -1 | 1 | 1 |
| 3 | Artzi | 2020 | Dev & Val | Retrospective | 417, 601 | 1 | 1 | 1 | 1 | 1 | 1 | 1 | 1 | 1 | 1 | 1 | 0 | 1 | 0 | 1 | 1 | 1 | 1 | -1 | 1 | 1 |
| 4 | Enquobahrie | 2015 | Dev | NCC | 358 | 1 | 1 | 1 | 1 | 1 | 1 | -1 | 1 | 1 | 1 | 1 | 1 | 1 | 0 | 1 | 1 | 1 | 1 | -1 | 1 | 1 |
| 5 | Amini | 2020 | Dev |  | 523 | 1 | 1 | 1 | 1 | 1 | 1 | -1 | 1 | 1 | 1 | 1 | 1 | 1 | 1 | 0 | 1 | 1 | 1 | -1 | 1 | 1 |
| 6 | Schaefer | 2018 | Dev | Prospective cohort | 8,381 | 1 | 1 | 1 | 1 | 1 | 1 | 1 | 1 | 1 | 1 | 1 | 1 | 1 | 0 | 1 | 0 | 1 | 1 | -1 | 1 | 1 |
| 7 | Hu | 2019 | Dev | Case-control | 2119 | 1 | 1 | 1 | 1 | 1 | 1 | 1 | 1 | 1 | 1 | 1 | 1 | 1 | 1 | -1 | 0 | 1 | 1 | -1 | 1 | 1 |
| 8 | Gao | 2020 | Dev |  | 19331 | 1 | 1 | 1 | 1 | 1 | 1 | 1 | 1 | 1 | 1 | 1 | 1 | 1 | 1 | 1 | 0 | 1 | 1 | -1 | 1 | 1 |
| 9 | Snyder | 2020 | Dev & Val | Cohort | 105379 | 1 | 1 | 1 | 1 | 1 | 1 | 1 | 1 | 1 | 1 | 1 | 1 | 1 | 0 | 1 | 0 | 1 | 1 | -1 | 1 | 1 |
| 10 | Jiang | 2019 | Dev |  | 431 | 1 | 1 | 1 | 1 | 1 | 1 | 1 | 1 | 1 | 1 | 1 | 1 | 1 | 1 | 0 | 1 | 1 | 1 | -1 | 1 | 1 |
| 11 | Liu | 2021 | Dev | Prospective cohort | 19,331 | 1 | 1 | 1 | 1 | 1 | 1 | 1 | 1 | 1 | 1 | 1 | 1 | 1 | 0 | 1 | 0 | 1 | 1 | 1 | 1 | 1 |
| 12 | Zheng | 2019 | Dev | Retrospective & Prospective | 4771 | 1 | 1 | 1 | 1 | 1 | 1 | 1 | 1 | 1 | 1 | 1 | 1 | 1 | 0 | -1 | 0 | 1 | 1 | -1 | 1 | 1 |
| 13 | White | 2016 | Dev | Prospective cohort | 1303 | 1 | 1 | 1 | 1 | 1 | 1 | 1 | 1 | 1 | 1 | 1 | 1 | 1 | 1 | 1 | 1 | 1 | 1 | -1 | 1 | 1 |
| 14 | Huang | 2020 | Dev | Prospective | 474 | 1 | 1 | 1 | 1 | 1 | 1 | 1 | 1 | 1 | 1 | 1 | 1 | 1 | 1 | 0 | 0 | 1 | 1 | -1 | 1 | 1 |
| 15 | Sebastien | 2015 | Dev | NCC | 792 | 1 | 1 | 1 | 1 | 1 | 1 | 1 | 1 | 1 | 1 | 1 | 1 | 1 | 1 | 1 | 0 | 1 | 1 | -1 | 1 | 1 |
| 16 | Harrison | 2015 | Dev | Retrospective cohort | 224 | 1 | 1 | 1 | 1 | 1 | 1 | 1 | 1 | 1 | 1 | 1 | 1 | 1 | 1 | -1 | 0 | 1 | 1 | -1 | 1 | 1 |
| 17 | Zhenyu | 2016 | Dev | Retrospective case-control | 836 | 1 | 1 | 1 | 1 | 1 | 1 | 1 | 1 | 1 | 1 | 1 | 1 | -1 | 1 | -1 | 0 | 1 | 1 | -1 | 1 | 1 |
| 18 | Harrison | 2018 | Dev |  | 103 | 1 | 1 | 1 | 1 | 1 | 1 | 1 | 1 | 1 | 1 | 1 | 1 | 1 | 1 | -1 | 1 | 1 | 1 | -1 | 1 | 1 |
| 19 | Wang | 2021 | Dev & Val | Retrospective cohort | 1640 | 1 | 1 | 1 | 1 | 1 | 1 | 1 | 1 | 1 | 1 | 1 | 1 | 1 | 1 | -1 | 0 | 1 | 1 | -1 | 1 | 1 |
| 20 | Zhang | 2020 | Dev |  | 1158 | 1 | 1 | 1 | 1 | 1 | 1 | 1 | 1 | 1 | 1 | 1 | 1 | 1 | 1 | -1 | 0 | 1 | 1 | -1 | 1 | 1 |

**Figure S2. The trend of published articles**

a. <14 weeks

<14 weeks

|  |  |
| --- | --- |
| 2010 | 1 |
| 2011 | 1 |
| 2013 | 1 |
| 2014 | 3 |
| 2015 | 3 |
| 2016 | 1 |
| 2017 | 5 |
| 2018 | 4 |
| 2019 | 2 |
| 2020 | 4 |
| 2021 | 6 |

b. 14-20 weeks

14-20 weeks

|  |  |
| --- | --- |
| 2015 | 4 |
| 2016 | 2 |
| 2018 | 2 |
| 2019 | 4 |
| 2020 | 6 |
| 2021 | 2 |
